# Spatial dynamics of SARS-Cov-2 and reduced risk of contagion: evidence from the second Italian epidemic wave^*^

**DOI:** 10.1101/2020.11.08.20227934

**Authors:** Paolo Buonanno, Sergio Galletta, Marcello Puca

## Abstract

We highlight a negative association between the severity of the first-wave of SARS-Cov-2 and the spread of the virus during the second-wave. Analyzing data of a sample of municipalities from the Italian region of Lombardy, we find that a one standard deviation increase in excess of mortality during the first-wave is associated with a reduction of approximately 30% in the number of detected infected individuals in the initial phase of the second-wave. Our findings may reflect a behavioral response in more severely hit areas as well as a cross-protection between successive waves.

**JEL Classification:** I10; I18

## 1. Introduction

The COVID-19 pandemic has already produced a broad negative effect around the world. As of the end of October, more than 50 million SARS-CoV-2 infections have been detected worldwide, while the number of reported victims passes the 1 million. The surge of new cases during the fall of 2020 in different parts of the globe – in particular many European countries – seems to confirm what most experts were predicting, the arriving of a second-wave of infections (Xu and Li, 2020). Recent contributions (Cacciapaglia et al., 2020; Fan, Yang, Lin, Zhao, Yang, and He, Fan et al.; Grech and Cuschieri, 2020; Leung et al., 2020; López and Rodó, 2020) forecast and projected the time evolution of COVID-19 pandemic across regions to provide quantitative evidence in support of policymaker and government decision to tackle the spread of the pandemic. Like other countries across the world, Italy, and in particular Lombardy, is experiencing a second wave. Figure 1 shows the first wave in Lombardy in March-May 2020 (spring wave) followed by a second and higher peak in October-November 2020 (fall wave).

**Figure 1:**
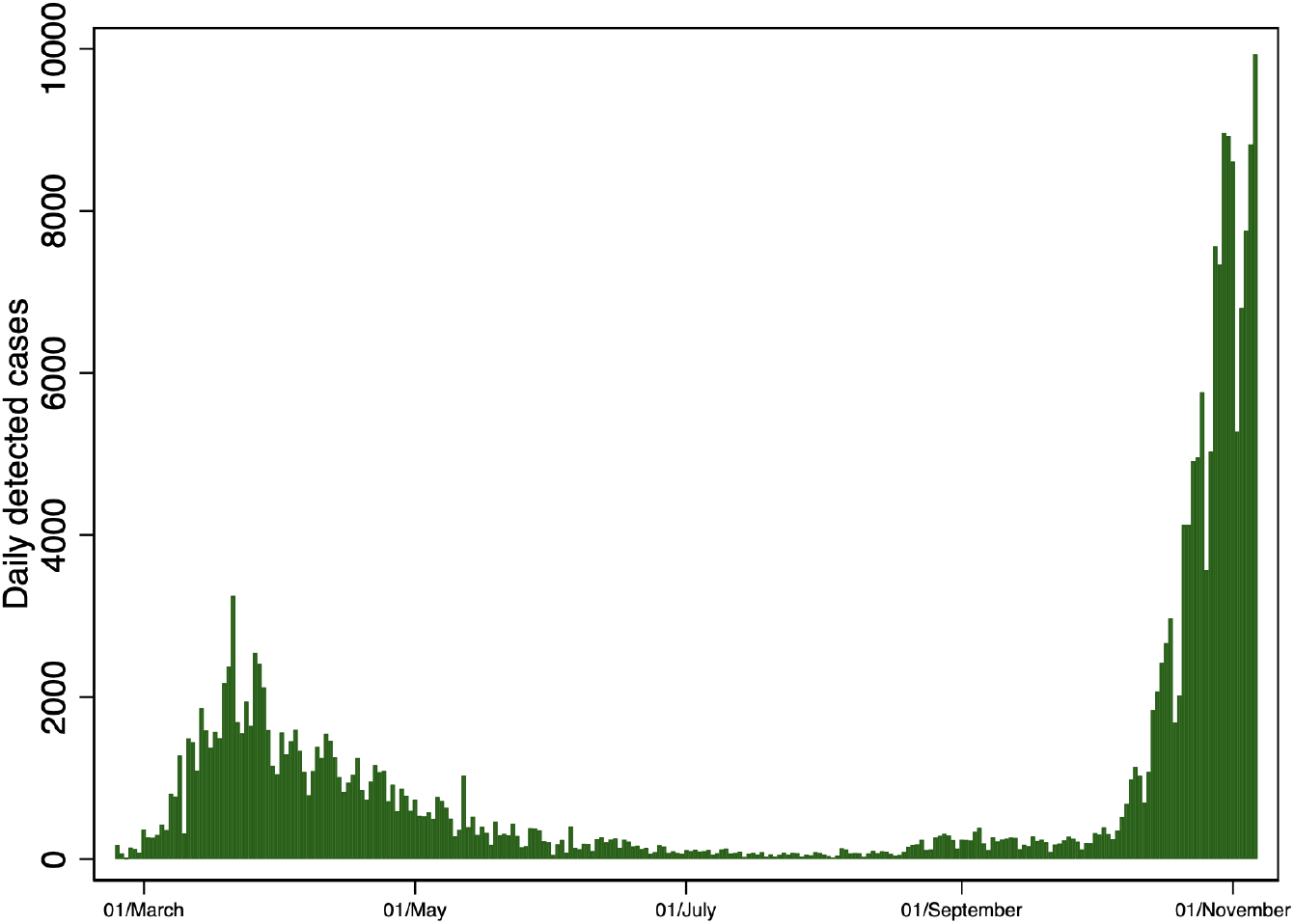
COVID-19 cases in Lombardy: first and second waves *Notes:* The figure shows the number of detected COVID-19 cases from February 21 to November 5, 2020.

The COVID-19 pandemic evolution presents some similarities with the 1918 Pandemic Influenza (Spanish Flu) that occurred in three waves from the spring of 1918 to the winter of 1919. As widely discussed in Barry (2004), the 1918 Pandemic Influenza has been characterized by a mild first wave in spring 1918, an extremely lethal second wave in fall 1918, followed by a less severe third wave in winter and spring 1919. The exposure to influenza in the spring of 1918 provided some immunization during the following waves. As noted in Barry et al. (2008) for the USA: “the intensity of the first wave may have differed across US cities and countries and may partly explain geographical variation in pandemic mortality rates in the fall”.

In this paper, we aim to study the relationship between the severity of the first-wave of SARS-Cov-2 occurred in Spring 2020 and the spread of the virus during the second-wave in Fall 2020. We focus on municipalities of the Italian region of Lombardy that has been severely hit during the first-wave and sadly known as the European epicenter. Exploiting both a traditional regression framework and a spatial econometric model to account for possible spatial effects and cross-municipalities interactions, we find that a one standard deviation increase in the excess of mortality during the first-wave is associated with a reduction of approximately 30% in the number of detected infected individuals. This could be due both to a behavioral response of individuals in the more severely hit areas and a higher immunization in the first wave that could contribute to a slower spread of the virus during the second wave of the epidemic.

Our results suggest that policymakers and health authorities should collaborate to design containment measures tailored only on small geographic entities and that should consider immune protection acquired during previous waves, despite herd immunity is far to be reached (Britton et al., 2020; Brett and Rohani, 2020). The rest of the paper is organized as follows. We describe data and the empirical methodology in section 2. Section 3 presents the estimation results, and section 5 concludes.

## 2. Data and Methods

### Data

We combine data from several sources. Specifically, we create a measure of excess mortality, which compares the number of deaths observed in Lombardy during the first COVID-19 wave (between January 1 and May 31, 2020), to the average number of deaths observed during the same time-span for all the years between 2015 and 2019. We gather mortality data released on October 22, 2020 from the Italian National Statistical Institute (ISTAT).^1^ The dataset includes the total mortality of 1,501 out of the 1,506 municipalities of Lombardy, virtually covering the entire regional population.

We merge this information with data on the number of cases observed during the more recent (and not yet ended) second COVID-19 wave, from September 1 to November 1, 2020.^2^ The dataset contains, for each Lombardy’s municipality, the number of new cases. We use these variables in section 3 to compare the number of COVID-19 cases detected during the second-wave with the number of the excess of deaths observed in the first-wave.

To clean our estimations from possible confounding factors, we also collect information on: (i) the age structure of the population and compute the share of individuals over 60 years (*Share of over 60*) from ISTAT; (ii) an indicator of the air quality (*Pollution*) based on the prevalence of fine particulate matters PM 2.5 over the period 2017 to 2019 (*Pollution*) collected from ARPA Lombardia (Regional Environmental Protection Agency); and (iii) a proxy for municipality civic capital, that is the share of recycling of urban solid waste (*Civic capital*) collected from ISTAT, which should capture the propensity of individuals to contribute to a public good and to comply with legal and social norms.

Table 1 reports basic summary statistics for the variables used in our analysis.

**Table 1:** Summary Statistics

| Variable | (1)<br>Mean | (2)<br>S.D. | (3)<br>Min | (4)<br>Max | (5)<br>N |
| --- | --- | --- | --- | --- | --- |
| Per-capita first-wave excess mortality | 0.003 | 0.004 | -0.022 | 0.034 | 1,501 |
| Per-capita second-wave cases | 0.007 | 0.009 | 0.000 | 0.271 | 1,501 |
| Total population | 6,725.063 | 37,524.981 | 30 | 1,396,059 | 1,501 |
| Population density | 572.035 | 812.655 | 0.000 | 7,771.109 | 1,499 |
| Share of over 60 | 28.786 | 5.211 | 11.939 | 58.262 | 1,489 |
| Pollution | 18.300 | 4.691 | 4.346 | 29.335 | 1,480 |
| Civic capital | 67.715 | 17.928 | 10.610 | 93.870 | 1,475 |

*Notes:* This table reports summary statistics of the variables used in our analysis.

### Methods

In section 3 we analyze our dataset using both ordinary least-squares (OLS) and generalised spatial two stage least squares (GS2SLS) estimator of Kelejian and Prucha (1998). Moreover, in our OLS regression we account for spatial correlation across nearby units using Conley standard errors (Conley, 1999). Specifically, we estimate both the following model

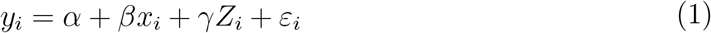

where *y* is the second-wave number of detected cases per 100k inhabitants, *x* is the excess mortality per 100k inhabitants observed during the first epidemic wave, *α* is the constant, *Z* is a set of control variables, and *ε* is a disturbance term. We are now ready to illustrate the results of our estimations.

## 3. Results

### Basics

We start our investigation analyzing the geographic distributions of the first-wave excess mortality and the second-wave number of cases. Figure 2 reports for each municipality of Lombardy, the first-wave excess of mortality per 100k inhabitants (left-hand-side panel) and the second-wave cumulative number of positive cases per 100k inhabitants (right-hand-side panel). Even a rough inspection of the maps reveals that the number of second-wave cases is lower in the municipalities that were more severely hit during the first epidemic wave, suggesting that the risk of contagion might be lower in those areas.

**Figure 2:**
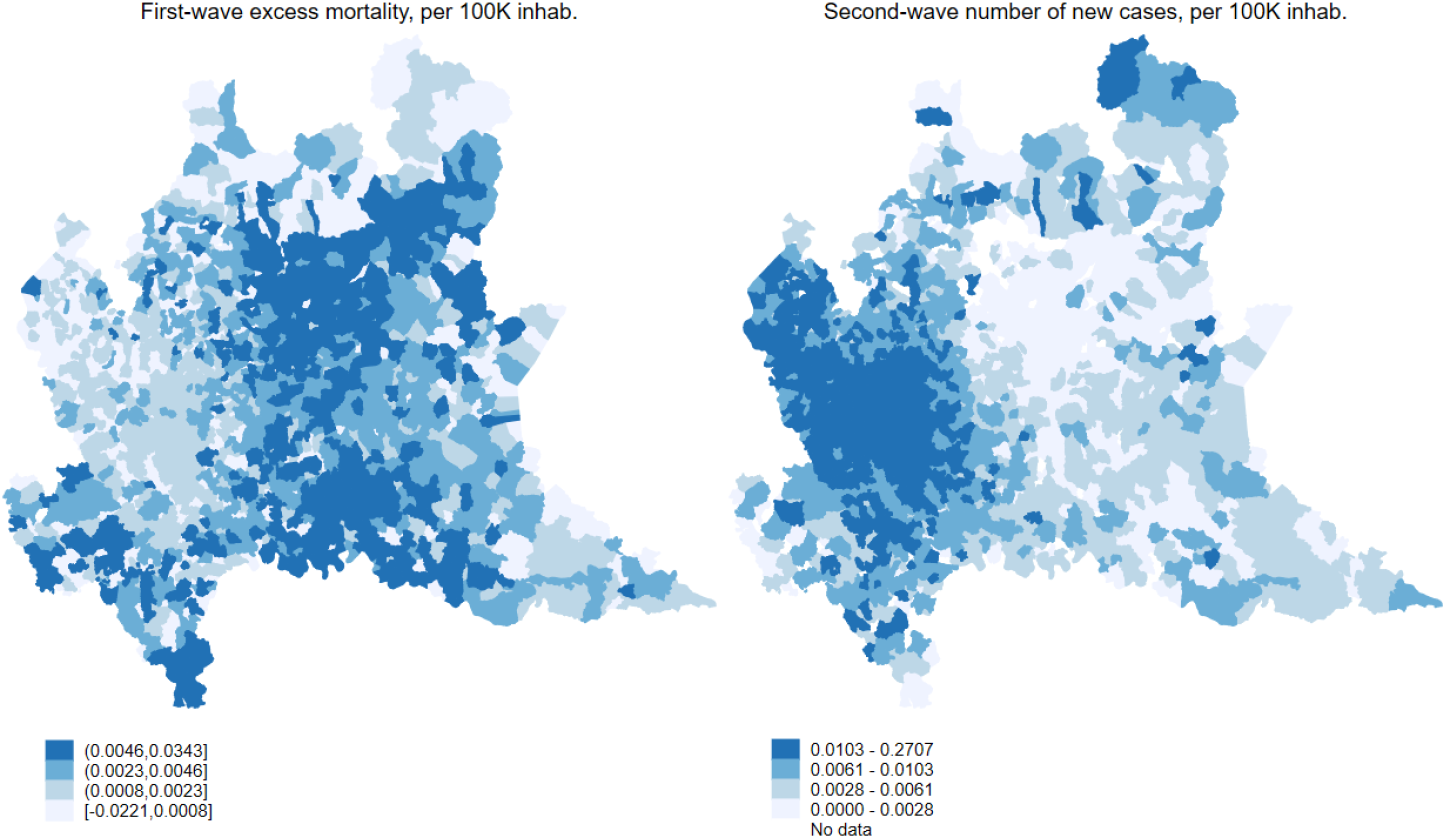
Geographic distribution of first vs second COVID19 waves *Notes:* The figure show the geographic distribution of the first-wave excess of mortality (left-hand-side panel) and the second-wave cumulative number of positive cases (right-hand-side panel).

To confirm our intuition, we perform a more in-depth empirical investigation with regression analysis. Table 2 reports ordinary least-squares regressions with the number of COVID-19 cases per 100k inhabitants detected during the second wave (*Second-wave cases*) as the dependent variable and the excess of mortality per 100k inhabitants observed during the first wave (*First-wave excess mortality*) as the main explanatory variable. The excess-mortality coefficient has a negative sign, and it is statistically significantly different from zero at 5% level. This relationship is robust to the inclusion of a minimal set of demographic controls such as population density (*Population density*), the share of individuals older than 60 (*Share of over 60*), a proxy of air pollution (*Pollution*), and a proxy of civic capital measured by the share of solid-waste recycling (*Civic capital*). We report in columns (1)-(5) robust standard errors and, in column (6), Conley corrected standard errors. The standardized coefficient of *First-wave excess mortality* suggests that an increase of one standard deviation of the first-wave excess mortality corresponds approximately to a 30% reduction of detected second-wave cases. Finally, we illustrate this negative relationship in Figure 3, which is the visual representation of the model (5) of Table 2, i.e., the binned scatter plot of a regression model including the whole set of control variables.

**Table 2:** OLS estimates

| Dep. Variable: Second-wave cases | (1) | (2) | (3) | (4) | (5) | (6) |
| --- | --- | --- | --- | --- | --- | --- |
| First-wave excess mortality | -0.6455**<br>(0.2531) | -0.5967**<br>(0.2544) | -0.5984**<br>(0.2621) | -0.6138**<br>(0.2679) | -0.6153**<br>(0.2704) | -0.6369**<br>(0.2781) |
| Population density |  | 0.2351***<br>(0.0177) | 0.2344***<br>(0.0167) | 0.2102***<br>(0.0205) | 0.2110***<br>(0.0206) | 0.2090***<br>(0.0369) |
| Share of over 60 |  |  | 0.0958<br>(4.9032) | 4.8741<br>(6.5172) | 5.1981<br>(6.9296) | 5.3448<br>(9.6926) |
| Pollution |  |  |  | 12.1499*<br>(6.5179) | 11.3421*<br>(5.9557) | 11.5993<br>(8.7915) |
| Civic capital |  |  |  |  | 0.3636<br>(1.1460) | 0.4580<br>(2.0478) |
| Observations | 1,501 | 1,499 | 1,489 | 1,480 | 1,465 | 1,454 |
| R<sup>2</sup> | 0.0892 | 0.1348 | 0.1347 | 0.1373 | 0.1375 | 0.1385 |
| S.E. | Robust | Robust | Robust | Robust | Robust | Conley |

*Notes:* This table reports ordinary least-squares regressions. The dependent variable is *Second-wave cases*, the COVID-19 cases per 100k inhabitants detected during the second wave. The main explanatory variable is *First-wave excess mortality* the excess of mortality observed during the first wave per 100k inhabitants. Controls include: (i) the share of individuals over 60 years (*Share of over 60*); (ii) an indicator of the air quality (*Pollution*) based on the prevalence of fine particulate matters PM 2.5 over the period 2017 to 2019 (*Pollution*) and (iii) a proxy of a municipality civic capital, that is the share of recycling of urban solid waste (*Civic capital*). Robust standard errors are presented in parentheses. \*, \*\* and \*\*\* denote rejection of the null hypothesis of the coefficient being equal to 0 at 10%, 5% and 1% significance level, respectively. In column 6, standard errors are computed using Conley standard errors (Conley, 1999).

**Figure 3:**
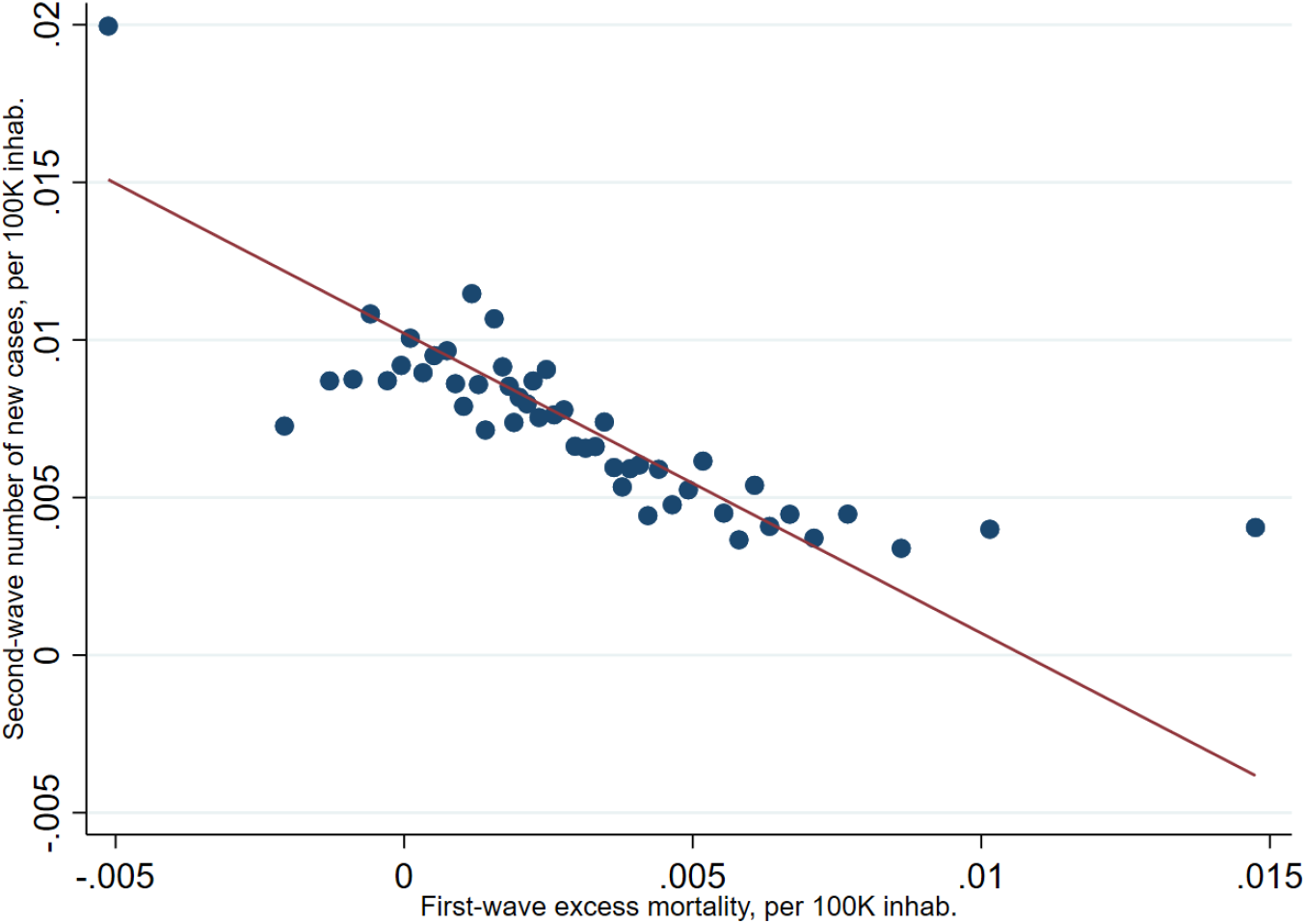
Bin scatter of main regression *Notes:* The figure shows the binned scatter plot of the regression model (5) of Table 2.

### Spatial dynamics

As an additional check we control for possible spatial effects and cross-municipalities interactions (e.g. Drukker et al., 2013). There is no reason to believe that infections and contagions follow the administrative boundaries of municipalities. Omitting to take them into account may reduce the efficiency of our estimates and bias them. To address this issue, we estimate a spatial model using the generalized spatial two-stage least squares (GS2SLS) estimator of Kelejian and Prucha (1998). We report in Table 3 the results of spatial estimations of the correlation between first-wave excess mortality and second-wave cases, which reproduces the same specification of column 5 of 2. We employ a contiguity matrix, and we implement a spatial autoregressive model (column (1)), a spatial error model (column (2)), and a model that combines the two by considering both a spatial lag and a spatial error structure (column (3)). Allowing for a spatial structure in our data does not alter our baseline estimates: the coefficient of *First-wave excess mortality* is negative and statistically significant at 1%.

**Table 3:** Spatial estimates

| Dep. Variable: Second-wave cases | (1) | (2) | (3) |
| --- | --- | --- | --- |
| First-wave excess mortality | -0.3328***<br>(0.0602) | -0.5374***<br>(0.0560) | -0.2715***<br>(0.0498) |
| Population density | 0.0007<br>(0.0359) | 0.1697***<br>(0.0355) | 0.0425**<br>(0.0193) |
| Share of over 60 | 4.3440<br>(4.8810) | 3.2409<br>(5.5031) | 3.7680<br>(2.8650) |
| Pollution | 3.9955<br>(5.9219) | 11.8042<br>(7.4300) | 3.0321<br>(2.9005) |
| Civic capital | 0.7890<br>(1.4141) | 0.7755<br>(1.6625) | 0.2302<br>(0.7439) |
| $\lambda$ | 0.8239***<br>(0.0859) | | 0.8118***<br>(0.0518) |
| $\rho$ | | 0.2574***<br>(0.0385) | -0.9480***<br>(0.1228) |
| Observations | 1,465 | 1,465 | 1,465 |

*Notes:* This table presents the results of a spatial model estimated using the generalized spatial two-stage least squares (GS2SLS) estimator of Kelejian and Prucha \(1998\). Included controls are the same as in the specification of column 5 of Table 2. We employ a contiguity matrix. A spatial lag model, a spatial error model, and a model that combines the two by considering both a spatial lag and a spatial error structure are respectively presented in columns (1), (2), and (3). $\lambda$ is the spatial lag term, while $\rho$ is the spatial error. The dependent variable is *Second-wave cases*, the COVID-19 cases per 100k inhabitants detected during the second wave. The main explanatory variable is *First-wave excess mortality* the excess of mortality observed during the first wave per 100k inhabitants. Controls include: (i) the share of individuals over 60 years (*Share of over 60*); (ii) an indicator of the air quality (*Pollution*) based on the prevalence of fine particulate matters PM 2.5 over the period 2017 to 2019 (*Pollution*) and (iii) a proxy of a municipality civic capital, that is the share of recycling of urban solid waste (*Civic capital*). Robust standard errors are presented in parentheses. \*, \*\* and \*\*\* denote rejection of the null hypothesis of the coefficient being equal to 0 at 10%, 5% and 1% significance level, respectively.

### The role of civic capital

Resent research has highlighted the role of civic capital in slowing down the spread of COVID-19 (Durante et al., 2020). To further explore such a phenomenon, we decompose the effect of *First-wave excess mortality* on *Second-wave cases* by interacting the former with the quartiles of *Civic capital*. While we omit the estimation results (available upon request) for brevity, we observe that the the reduction in the number of cases is significantly stronger for municipalities with higher levels of civic capital. Specifically, the coefficients of the interactions between *Second-wave cases* and the quartiles of *Civic capital* are all negative, and statistically significantly different from zero at 1% level. These results are consistent with the idea that civic-minded individuals internalize more than others the effect of their behavior on the spread of an infectious disease.

## 4. Discussion and policy implications

Two potentially simultaneous effects could explain the observed negative association between the first-wave severity and the current second-wave reduced risk of contagion. On the one hand, people living in areas that were severely affected during the first-wave are likely to have changed their behavior in a way that would make less likely the spread of the virus (e.g., more accurate use of protecting devices, more frequent sanitizing procedures, etc.). This might not be true in areas where, instead, many individuals did not have actual direct or indirect exposure to the consequences of the virus (e.g., a dead friend or relative). However, since the inclusion of a proxy of civic capital as a control variable did not affect our estimates, the observed heterogeneity could be explained by alternative mechanisms.

On the other hand, severe exposure to the virus during the first-wave may have immunized most of the population, contributing to a much slower spread of the virus during the second wave of the epidemic. As reported in previous work (Buonanno et al., 2020; Perico et al., 2020) certain provinces in Lombardy reported a very high level of predicted infections (considering a hypothetical fatality rate of 1%, the share of the population infected is estimated to be: 40% in the province of Bergamo, 36% in Cremona, 29% in Lodi and 19% in Brescia).

Our results show that policymakers interested in trading off the benefit of containment measures with the cost of shutting down economic activities should therefore tailor their policies to relatively small geographical areas.

Despite a robust and significant negative correlation between the first-wave excess mortality and second-wave contagions, we cannot convincingly disentangle whether the effect is due to a sort of herd immunity or to a behavioral response of individuals. Most likely, the two effects are simultaneous and complement each other.

## 5. Concluding remarks

We analyzed the correlation between the exposure to the first-wave COVID-19 pandemic in Lombardy and the current second-wave risk of contagion. We report that municipalities more severely hit by the epidemic during the first wave are experiencing fewer cases during the second wave. Although the results should be interpreted cautiously in light of the assumptions and limitations inherent to our approach, our results suggest that policymakers and health authorities should collaborate to design containment measures that are tailored only on small geographic entities.

## Data Availability

All data are publicly available.

https://www.istat.it/it/archivio/240401

https://datawrapper.dwcdn.net/iMArO/10/

https://www.datawrapper.de/_/567DW/

## Footnotes

1 See https://www.istat.it/it/archivio/240401 for more information

2 We obtained data from the Lombardy Region, which are available at https://datawrapper.dwcdn.net/iMArO/10/ and https://www.datawrapper.de/_/567DW/

## References

Barry, J. M. (2004). The great influenza: the epic story of the deadliest plague in history. New York: Viking Penguin.

Barry, J. M., C. Viboud, and L. Simonsen (2008). Cross-Protection between Successive Waves of the 1918–1919 Influenza Pandemic: Epidemiological Evidence from US Army Camps and from Britain. The Journal of Infectious Diseases 198 (10), 1427–1434.

Brett, T. S. and P. Rohani (2020). Transmission dynamics reveal the impracticality of COVID-19 herd immunity strategies. Proceedings of the National Academy of Sciences 117 (41), 25897–25903.

Britton, T., F. Ball, and P. Trapman (2020). A mathematical model reveals the influence of population heterogeneity on herd immunity to SARS-CoV-2. Science 369 (6505), 846–849.

Buonanno, P., S. Galletta, and M. Puca (2020). Estimating the severity of COVID-19: Evidence from the italian epicenter. PLoS ONE 15 (10).

Cacciapaglia, G., C. Cot, and F. Sannino (2020). Second wave covid-19 pandemics in europe: a temporal playbook. Scientific Reports 10 (1), 15514.

Conley, T. G. (1999). GMM estimation with cross sectional dependence. Journal of econometrics 92 (1), 1–45.

Drukker, D. M., P. Egger, and I. R. Prucha (2013). On two-step estimation of a spatial autoregressive model with autoregressive disturbances and endogenous regressors. Econometric Reviews 32 (5-6), 686–733.

Durante, R., L. Guiso, and G. Gulino (2020). Asocial capital: Civic culture and social distancing during covid-19. CEPR Discussion Paper (DP14820).

Fan, G., Z. Yang, Q. Lin, S. Zhao, L. Yang, and D. He. Decreased case fatality rate of covid-19 in the second wave: A study in 53 countries or regions. Transboundary and Emerging Diseases.

Grech, V. and S. Cuschieri (2020). COVID-19: A global and continental overview of the second wave and its (relatively) attenuated case fatality ratio. Early Human Development, 105211.

Kelejian, H. and I. Prucha (1998). A generalized spatial two-stage least squares procedure for estimating a spatial autoregressive model with autoregressive disturbances. Journal of Real Estate Finance and Economics 17 (1), 92–121.

Leung, K., J. T. Wu, D. Liu, and G. M. Leung (2020). First-wave covid-19 transmissibility and severity in china outside hubei after control measures, and second-wave scenario planning: a modelling impact assessment. The Lancet 395 (10233), 1382– 1393.

López, L. and X. Rodó (2020). The end of social confinement and covid-19 re-emergence risk. Nature Human Behaviour 4 (7), 746–755.

Perico, L., S. Tomasoni, T. Peracchi, A. Perna, A. Pezzotta, G. Remuzzi, and A. Benigni (2020). SARS-CoV-2 and Lombardy: Testing the impact of the first wave of the pandemic. EBioMedicine 61, 103069.

Xu, S. and Y. Li (2020). Beware of the second wave of COVID-19. The Lancet 395 (10233), 1321–1322.

